# Real-World Effectiveness of Nirmatrelvir/Ritonavir in Preventing Hospitalization Among Patients With COVID-19 at High Risk for Severe Disease in the United States: A Nationwide Population-Based Cohort Study

**DOI:** 10.1101/2022.09.13.22279908

**Authors:** Xiaofeng Zhou, Scott P. Kelly, Caihua Liang, Ling Li, Rongjun Shen, Heidi K. Leister-Tebbe, Steven G. Terra, Michael Gaffney, Leo Russo

**Affiliations:** Global Medical Epidemiology, Worldwide Medical and Safety, Pfizer Inc, New York, NY, USA; Global Product Development, Pfizer Inc, Collegeville, PA, USA; Global Product Development, Pfizer Inc, Andover, MA, USA; Statistical Research, Pfizer Inc, New York, NY, USA; Global Medical Epidemiology, Worldwide Medical and Safety, Pfizer Inc, Collegeville, PA, USA

## Abstract

**Objectives:** The aim of this analysis was to describe nirmatrelvir/ritonavir real-world effectiveness in preventing hospitalization among high-risk US COVID-19 patients during SARS-CoV-2 Omicron predominance.

**Design:** An ongoing population-based cohort study with retrospective and prospective collection of electronic healthcare data in the United States.

**Methods:** Data for this analysis were collected from the US Optum® de-identified COVID-19 Electronic Health Record (EHR) dataset during December 22, 2021−June 8, 2022. Key eligibility criteria for inclusion in the database analysis were ≥12-years-old; positive SARS-CoV-2 test, COVID-19 diagnosis, or nirmatrelvir/ritonavir prescription; and high risk of severe COVID-19 based on demographic/clinical characteristics. Potential confounders between groups were balanced using propensity score matching (PSM). Immortal time bias was addressed.

**Outcome measures:** Hospitalization rates within 30 (primary analysis) or 15 (sensitivity analysis) days from COVID-19 diagnosis overall and within subgroups were evaluated.

**Results:** Before PSM, the nirmatrelvir/ritonavir group (n=2811) was less racially diverse, older, and had higher COVID-19 vaccination rates and a greater number of comorbidities than the non-nirmatrelvir/ritonavir group (n=194,542). Baseline characteristics were well balanced across groups (n=2808 and n=10,849, respectively) after PSM. Incidence of hospitalization (95% CI) within 30 days was 1.21% (0.84%−1.69%) for the nirmatrelvir/ritonavir group and 6.94% (6.03%−7.94%) for the non-nirmatrelvir/ritonavir group, with a hazard ratio (95% CI) of 0.16 (0.11−0.22; 84% relative risk reduction). Incidence within 15 days was 0.78% (0.49%−1.18%) for the nirmatrelvir/ritonavir group and 6.54% (5.65%−7.52%) for the non-nirmatrelvir/ritonavir group; hazard ratio 0.11 (0.07−0.17; 89% relative risk reduction). Nirmatrelvir/ritonavir was effective in African American patients (hazard ratio, 0.35 [0.15−0.83]; 65% relative risk reduction). Relative risk reductions were comparable with overall results across ages and among vaccinated patients.

**Conclusions:** Real-world nirmatrelvir/ritonavir effectiveness against hospitalization during the Omicron era supports EPIC-HR efficacy among high-risk patients. Future research should confirm these early real-world results and address limitations.

## Introduction

COVID-19 remains a significant threat to global health.^1, 2^ Nirmatrelvir is a potent, selective SARS-CoV-2 main protease inhibitor that addressed the urgent need for orally administered SARS-CoV-2 antiviral agents for delivery outside hospital settings.^3, 4^ Nirmatrelvir is administered with the pharmacokinetic enhancer ritonavir (nirmatrelvir/ritonavir; Paxlovid) to prevent CYP3A4-associated degradation.^5^

The pivotal phase 3 EPIC-HR randomized trial evaluated nirmatrelvir/ritonavir efficacy and safety among non-hospitalized patients with COVID-19 at increased severe disease risk.^4^ Among patients who commenced nirmatrelvir/ritonavir treatment within 3 and 5 days of symptom onset, the relative risk of the composite measure of COVID-19−related hospitalization or all-cause death within 28 days was reduced by 88.9% and 87.8%, respectively.^4^ Nirmatrelvir/ritonavir was granted emergency use authorization (EUA) by the FDA in December 2021 for treatment of mild-to-moderate COVID-19 in adults and pediatric (≥12-years-old; ≥40 kg) patients at high risk for progression to severe disease.^5^

Real-world data are crucial for ascertaining treatment effectiveness in clinical practice, yet adjustment for confounding is critical.^6-8^ Patients who received COVID-19 vaccine or had confirmed previous SARS-CoV-2 infection were excluded from EPIC-HR, representing a difference between EPIC-HR patients and high-risk patients who may receive nirmatrelvir/ritonavir in clinical practice.^4^ Furthermore, data evaluating clinical effectiveness of nirmatrelvir/ritonavir against the Omicron variant, which emerged after EPIC-HR completion, are limited and may differ compared with efficacy data from the trial.^9-11^

This is the first nirmatrelvir/ritonavir real-world US-wide study assessing nirmatrelvir/ritonavir effectiveness at preventing hospitalization in high-risk COVID-19 patients during Omicron predominance. As African American individuals have disproportionately poor COVID-19 outcomes,^12^ we also examined nirmatrelvir/ritonavir effectiveness in this vulnerable population and other key patient subgroups.

## Methods

### Data Source

This study involved data analysis from ongoing data collection from electronic health records (EHRs) of US patients. Data were derived from the Optum® de-identified COVID-19 Electronic Health Record dataset (**Supplementary Appendix**), which includes patients who had documented clinical care from January 2007 through the most current monthly data release, a documented exposure to or testing for SARS-CoV-2 (positive or negative result) and a COVID-19 or acute respiratory illness diagnosis after February 2020. Patient demographics, mortality, and clinical interventions (eg, medications prescribed) collected from EHRs derived from both acute inpatient stays and outpatient visits are included. As of June 8, 2022, which includes the Omicron-predominant period, the COVID-19 database included ∼12 million individuals, derived from Optum’s EHR repository with >700 hospitals and 7000 clinics from all US states.^13^

### Study Design, Population, Definitions

Study population inclusion and exclusion criteria for this retrospective population-based cohort study are outlined in **Figure 1A**. Briefly, ≥12-year-old patients irrespective of COVID-19 vaccination status were included if they had a positive SARS-CoV-2 PCR/antigen test, COVID-19 diagnosis (ICD-10 U07.1), or were prescribed nirmatrelvir/ritonavir between December 22, 2021 (EUA approval date) and May 8, 2022.

**Figure 1.**
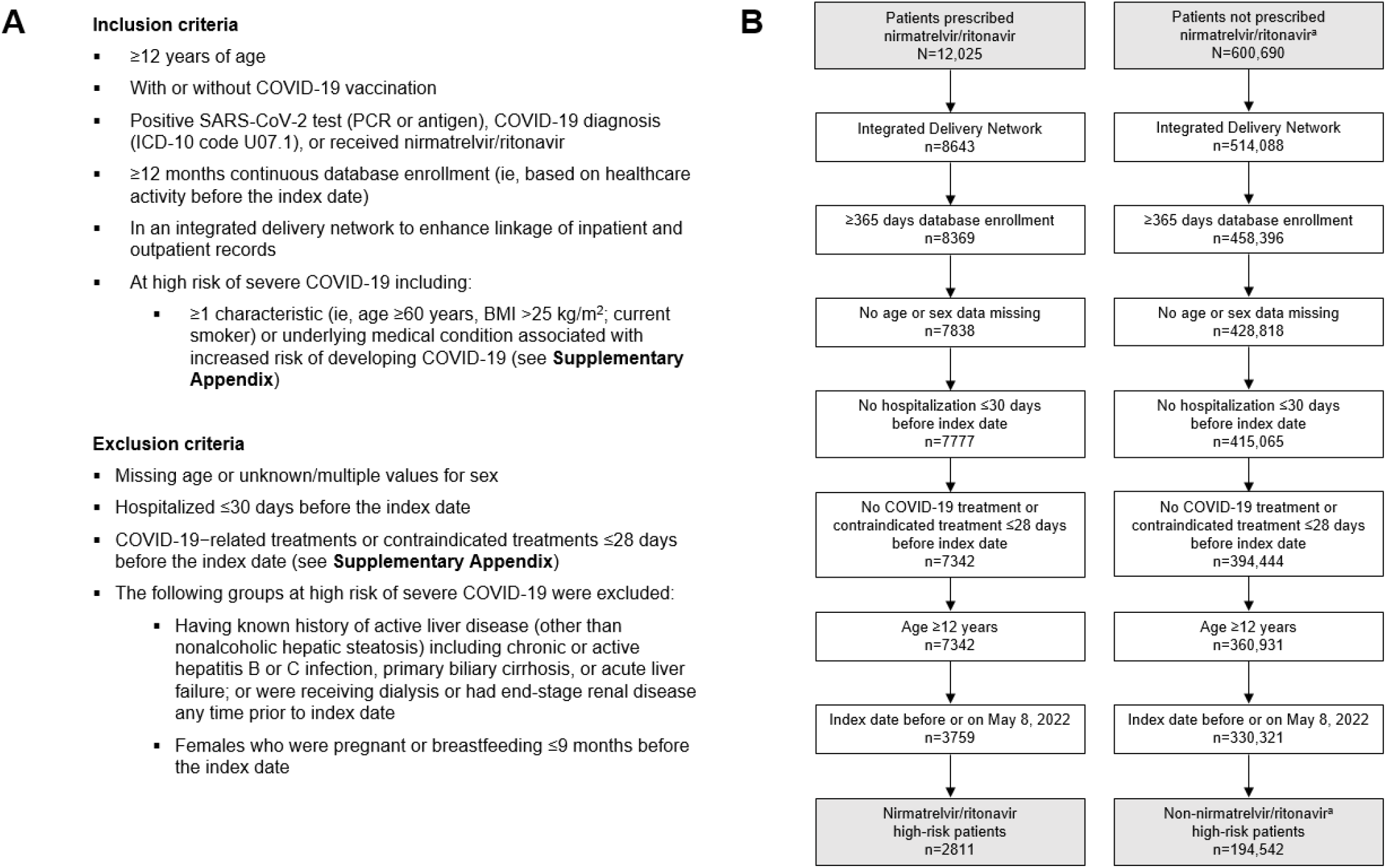
(A) Study inclusion and exclusion criteria and (B) patient attrition. ^a^Patients in the non-nirmatrelvir/ritonavir group were also not prescribed molnupiravir. BMI=body mass index.

Patients also had to have ≥1 characteristic or underlying medical condition associated with increased risk of developing severe COVID-19 in the year before cohort entry (**Supplementary Appendix**),^14-16^ which aligned closely with EPIC-HR.^4^ The nirmatrelvir/ritonavir group included all patients meeting inclusion criteria who were prescribed nirmatrelvir/ritonavir; the non-nirmatrelvir/ritonavir group included those not prescribed nirmatrelvir/ritonavir or molnupiravir.

The index date in the nirmatrelvir/ritonavir group was the date of first prescription. To avoid immortal time bias,^17^ index dates in the non-nirmatrelvir/ritonavir group were determined using the Prescription Time Distribution Method (PTDM)^18^ based on distribution of nirmatrelvir/ritonavir index dates relative to COVID-19 confirmation date (**Supplementary Appendix**). The baseline period was defined as 1 year before index date.

### Study Outcomes

All-cause hospitalization within 30 days of index date was used as the proxy of COVID-19−associated hospitalization since the study database cannot identify cause of hospitalization. A sensitivity analysis applied a 15-day risk window.

### Analyses

Continuous variables were described by means with standard deviations or medians with interquartile ranges. Dichotomous or categorical variables are presented using numbers and percentages. Conditions were captured using ICD-10 codes; treatments using National Drug Code and Healthcare Common Procedure Coding System codes; and procedures using ICD-10 procedure codes and CPT codes. Follow-up started at index date and extended to 30 days (primary analysis) or 15 days (sensitivity analysis), the first hospitalization occurrence, loss to follow-up, death, or data cutoff (June 8, 2022), whichever occurred first. A death date algorithm and lost to follow-up definition are presented in the **Supplementary Appendix**. All analyses were executed using SAS version 9.4.

#### Propensity Score Matching

Potential confounders between the nirmatrelvir/ritonavir and non-nirmatrelvir/ritonavir groups were adjusted using propensity score (PS) matching (PSM).^19^ Each patient in the nirmatrelvir/ritonavir group was matched with ≤4 non-nirmatrelvir/ritonavir patients on the basis of the PS obtained by logistic regression on prespecified baseline covariates including demographics (age, sex, race, ethnicity, region), insurance type, cohort entry month, healthcare utilization, COVID-19 vaccination status, number of past SARS-CoV-2 infections, comorbidities, and medical histories with absolute standardized differences >0.1, plus the 50 most common diagnoses, procedures, and prescriptions retained via stepwise selection before PSM. All prespecified covariates were at baseline other than COVID-19 vaccine receipt (ie, receiving ≥1 vaccine dose since December 13, 2020), number of past SARS-CoV-2 infections (since January 1, 2020), and age (at index date). After PSM, balance for each covariate was assessed using absolute standardized differences, where a difference of ≤0.1 was selected as the threshold for good balance^20^ between the nirmatrelvir/ritonavir and non-nirmatrelvir/ritonavir groups. The matching procedure utilized nearest neighbor matching with a caliper of 0.25 × standard deviation of the PS logit.

#### Outcomes Analysis

Hospitalization incidence proportion, incidence rate per 100 person-days, and respective 95% CIs were calculated. Hazard ratios (nirmatrelvir/ritonavir to non-nirmatrelvir/ritonavir) and 95% CIs were calculated using the Cox proportional hazards regression model conditional on the matching ratio. Any unbalanced variables after PSM were included in the final model for further adjustment. Relative risk reduction was derived by subtracting the hazard ratios from 1. Outcomes were also evaluated within subgroups defined by race (White, African American) and age (<65-year-olds, ≥65-year-olds), and in COVID-19 vaccine (ie, ≥1 dose) recipients.

## Results

### Patients

Among patients in the Optum COVID-19 database between December 22, 2021 and June 8, 2022, 12,025 patients were prescribed nirmatrelvir/ritonavir and 600,690 patients with positive SARS-CoV-2 tests or COVID-19 diagnoses were not prescribed nirmatrelvir/ritonavir (**Figure 1B**). After applying inclusion and exclusion criteria, 2811 and 194,542 patients were included in the nirmatrelvir/ritonavir and non-nirmatrelvir/ritonavir groups, respectively.

Before PSM, differences in characteristics between patients in the nirmatrelvir/ritonavir versus non-nirmatrelvir/ritonavir groups emerged, with patients in the nirmatrelvir/ritonavir group being less racially diverse; older; using more Medicare and less Medicaid; having a greater number of comorbidities; and having higher COVID-19 vaccination rates (**Table 1**). After PSM, 2808 and 10,849 patients remained in the nirmatrelvir/ritonavir and non-nirmatrelvir/ritonavir groups, respectively, and patients were well-matched regarding demographic and baseline clinical characteristics **(Table 1; Figure 2)**. Across groups after PSM, median age was 62.0−63.0 years, 41.8%−42.1% of patients were male, and 66%−68% were vaccinated. Healthcare utilization is shown in **Table S1**. Hospitalizations by month are shown in **Figure S1**. Death within 30 days of index date occurred in 7 (0.25%) and 100 (0.92%) patients in the nirmatrelvir/ritonavir and non-nirmatrelvir/ritonavir groups, respectively. Death within 15 days occurred in 5 (0.18%) nirmatrelvir/ritonavir patients and 78 (0.72%) non-nirmatrelvir/ritonavir patients. Percentages lost to follow-up are presented in **Supplementary Appendix**.

**Table 1.**
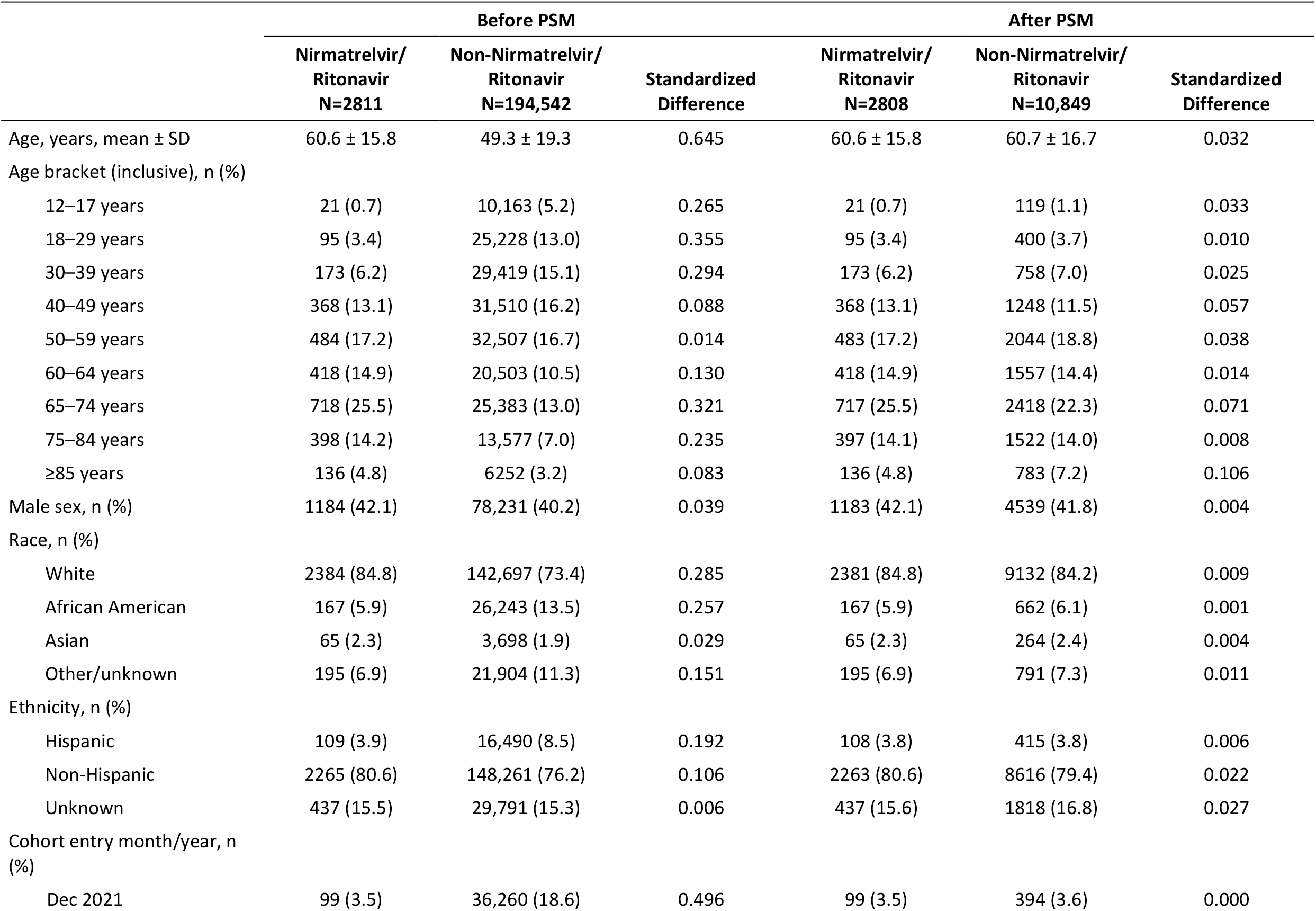
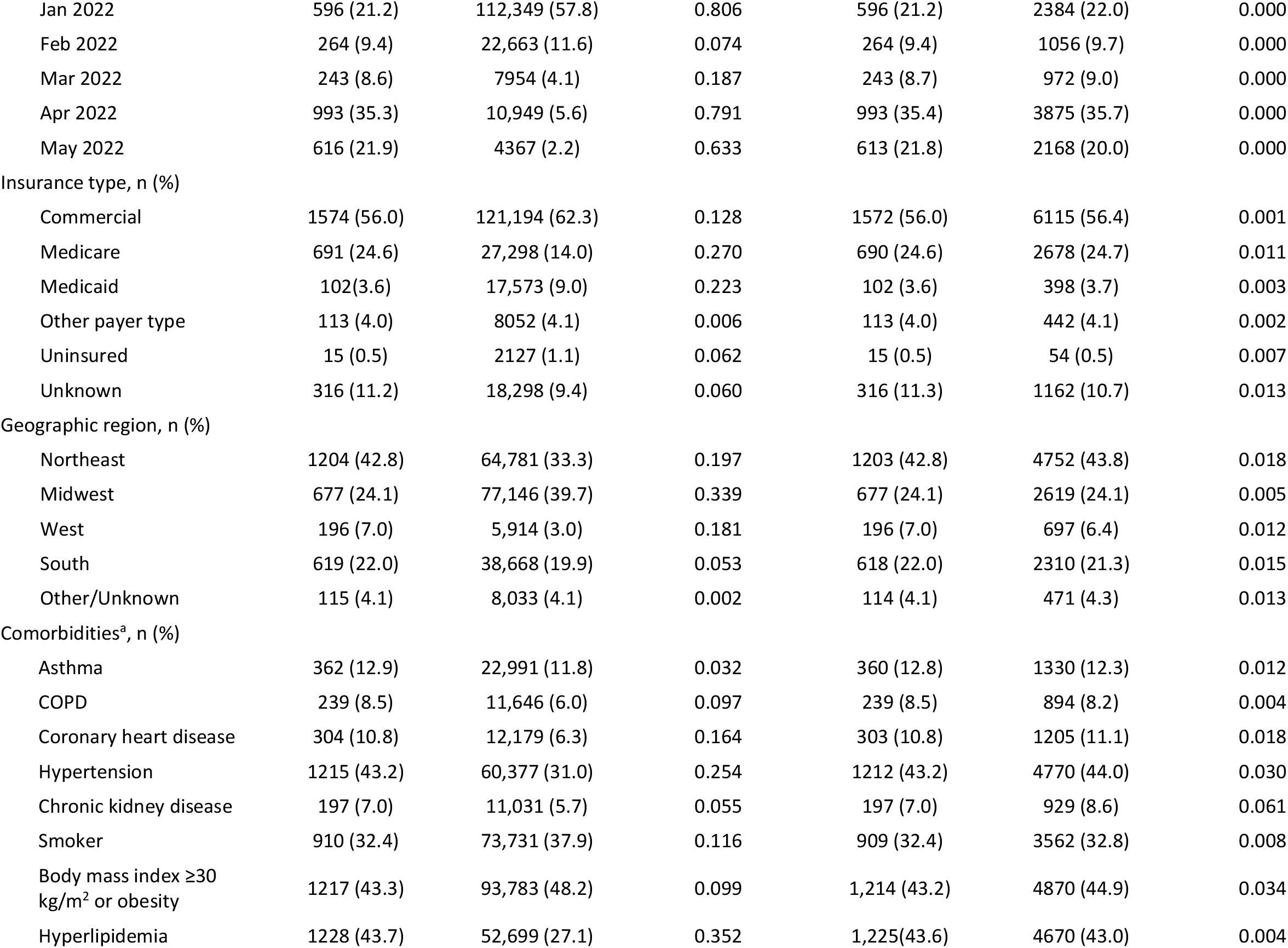
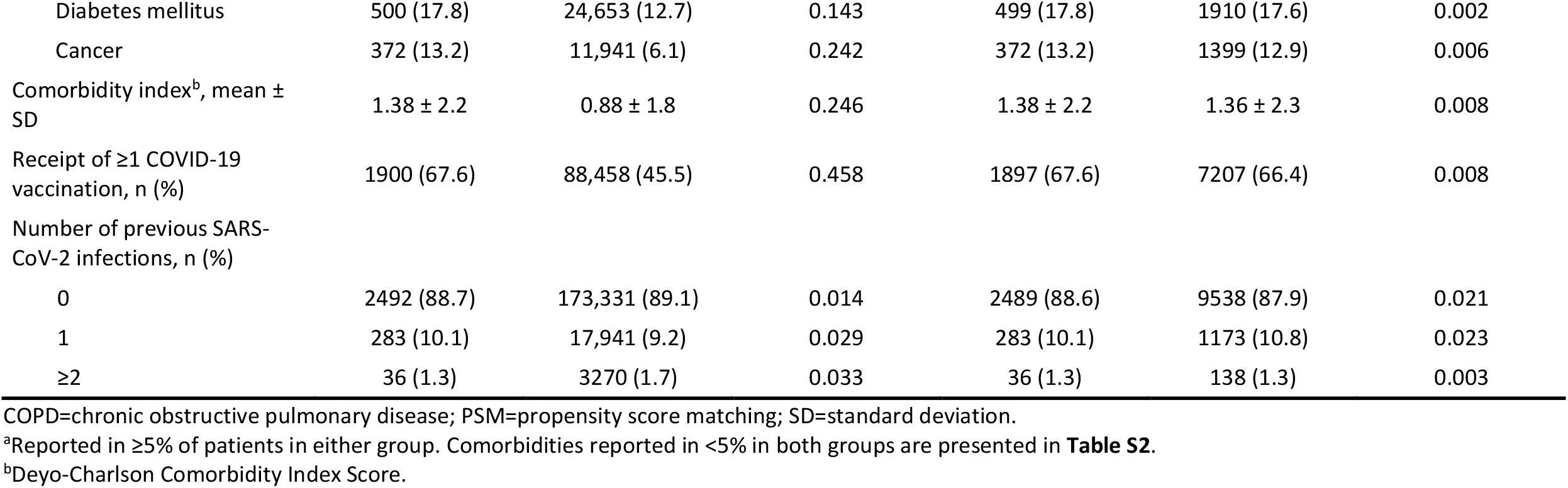
Baseline demographics and clinical characteristics before and after PSM.

**Figure 2.**
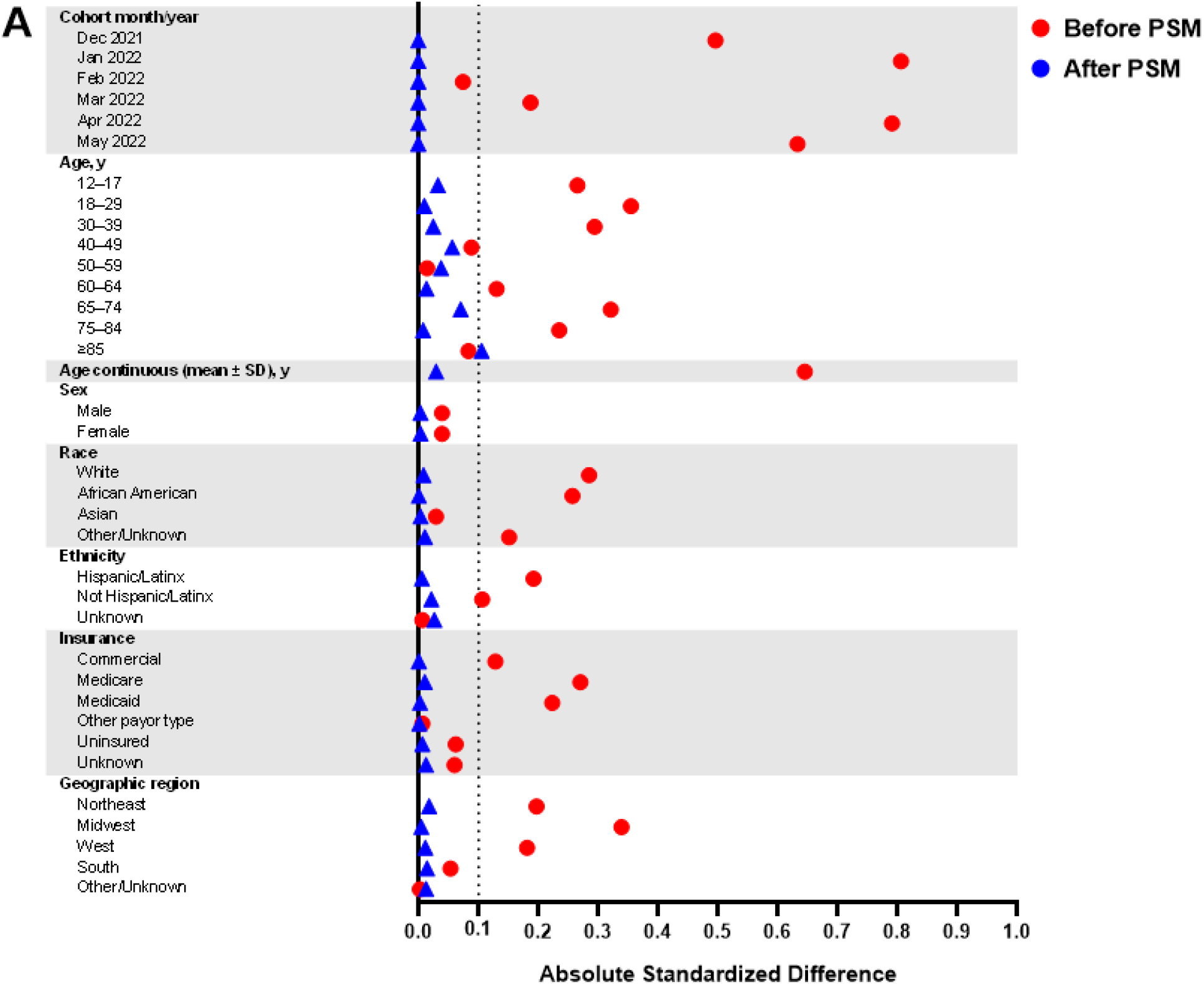
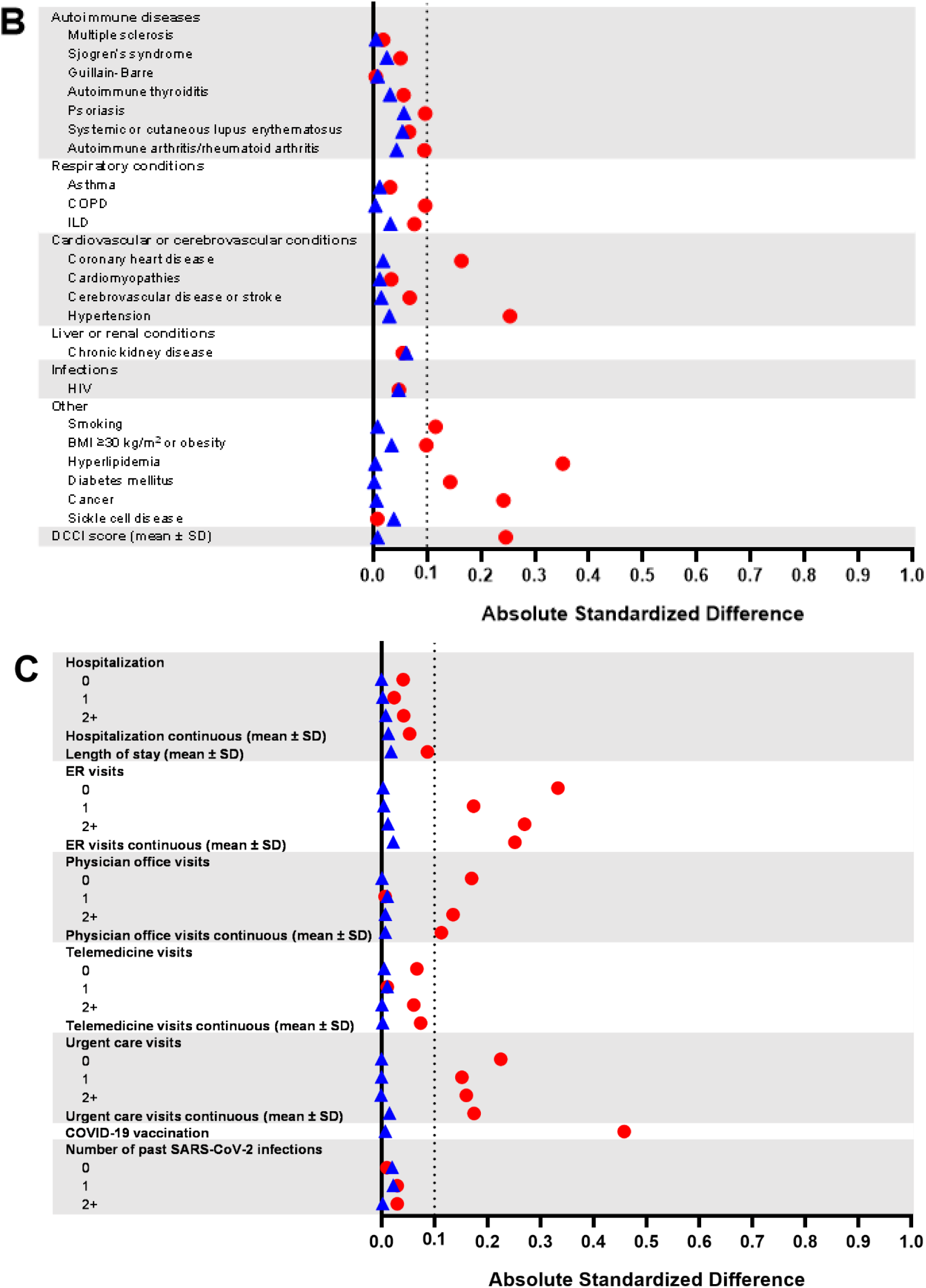
Standardized differences in baseline (A) demographics, (B) comorbidities, and (C) healthcare utilization, COVID-19 vaccination, and prior COVID-19 infection before and after PSM. The dotted line indicates a standardized difference of 0.1, which was selected as the threshold for good balance between the nirmatrelvir/ritonavir and non-nirmatrelvir/ritonavir groups. For before PSM, nirmatrelvir/ritonavir, n=2811; non-nirmatrelvir/ritonavir, n=194,542. For after PSM, nirmatrelvir/ritonavir, n=2808; non-nirmatrelvir/ritonavir, n=10,849. BMI=body mass index; COPD=chronic obstructive pulmonary disease; DCCI=Deyo-Charlson Comorbidity Index; ER=emergency room; ILD=interstitial lung disease; PSM=propensity score matching; SD=standard deviation.

### Effectiveness

#### Hospitalization

Among the high-risk population included after PSM, 34 and 752 patients in the nirmatrelvir/ritonavir and non-nirmatrelvir/ritonavir groups, respectively, were hospitalized within 30 days of index date (**Table 2**). Incidence (95% CI) was 1.21% (0.84%‒1.69%) and 6.94% (6.03%‒7.94%), with a hazard ratio (95% CI) of 0.16 (0.11‒0.22; 84% relative risk reduction) in the nirmatrelvir/ritonavir group. In the sensitivity analysis, 22 and 708 patients, respectively, were hospitalized within 15 days of index date, corresponding to an incidence of 0.78% (0.49%−1.18%) and 6.54% (5.65%−7.52%), with a hazard ratio of 0.11 (0.07‒0.17; ie, 89% relative risk reduction).

**Table 2.** Hospitalization among patients with COVID-19 by nirmatrelvir/ritonavir treatment status after PSM.

|  | **Nirmatrelvir/Ritonavir** | **Non-Nirmatrelvir/Ritonavir** |
| --- | --- | --- |
| **Hospitalization within 30 days** |  |  |
| Patients at risk, n | 2808 | 10,849 |
| Events, n | 34 | 752 |
| Incidence proportion, % (95% CI) | 1.21 (0.84, 1.69) | 6.94 (6.03, 7.94) |
| Person-days | 55,961 | 204,980 |
| Incidence rate, per 100 person-days (95% CI) | 0.06 (0.04, 0.08) | 0.37 (0.32, 0.43) |
| Hazard ratio (95% CI) | 0.16 (0.11, 0.22) | - |
| Relative risk reduction, % (95% CI) | 84 (78, 89) | - |
| **Hospitalization within 15 days** |  |  |
| Patients at risk, n | 2808 | 10,849 |
| Events, n | 22 | 708 |
| Incidence proportion, % (95% CI) | 0.78 (0.49, 1.18) | 6.54 (5.65, 7.52) |
| Person-days | 31,721 | 115,096 |
| Incidence rate, per 100 person-days | 0.07 (0.04, 0.11) | 0.62 (0.53, 0.71) |
| Hazard ratio (95% CI) | 0.11 (0.07, 0.17) | - |
| Relative risk reduction, % (95% CI) | 89 (83, 93) | - |

PSM=propensity score matching.
Hazard ratios and 95% CIs were computed using Cox proportional hazards regressions models conditional on the matching ratio. All covariates were well balanced, so no adjustment was needed in final model. Relative risk reductions were derived by subtracting the hazard ratios from 1.

#### Subgroup Analyses

In the non-nirmatrelvir/ritonavir group, the 30-day hospitalization incidence (95% CI) after PSM was higher in African American patients compared with White patients (8.76% [6.72−11.18] vs 6.94% [6.43−7.48]). In the nirmatrelvir/ritonavir group, hospitalization incidence was reduced in both racial groups, but the disparity between African American and White patients increased (4.79% [2.09−9.22] vs 1.01% [0.65−1.5]). Nirmatrelvir/ritonavir hazard ratios (95% CIs) for 30-day hospitalization were 0.14 (0.09−0.21; ie, 86% relative risk reduction) for White patients and 0.35 (0.15−0.83; ie, 65% relative risk reduction) for African American patients (**Figure 3A, Table S3**). Reductions in hospitalization risks in ≥65-year-old and <65-year-old patients and in vaccinated patients were consistent with overall findings (**Figure 3A−B**).

**Figure 3.**
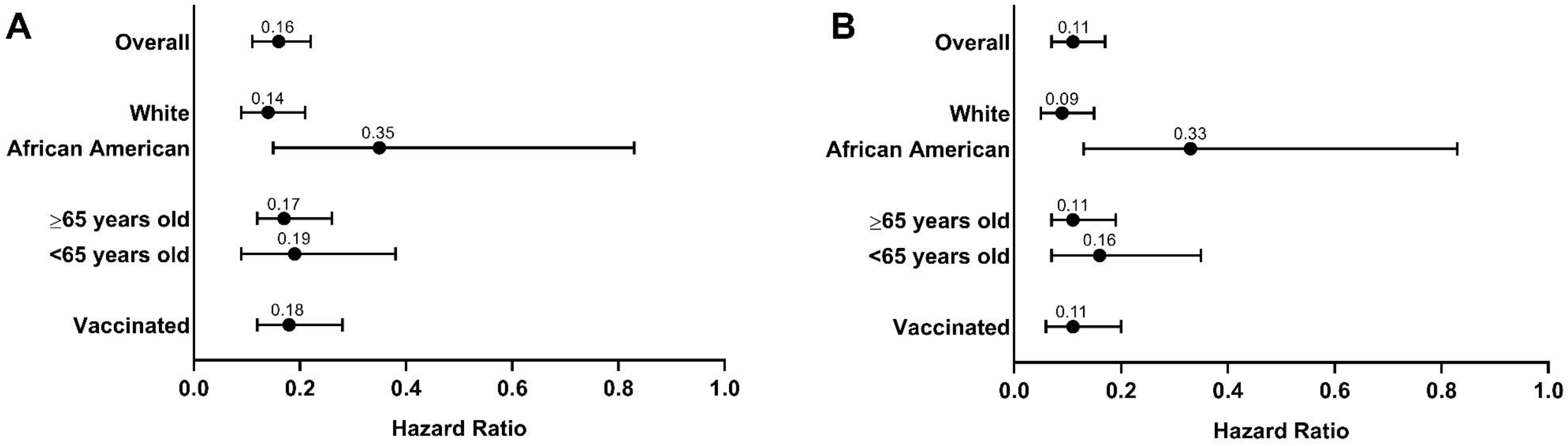
Hazard ratios for hospitalization within (A) 30 and (B) 15 days among subgroups of patients with COVID-19 after PSM. Hazard ratios and 95% CIs were computed using Cox proportional hazards regressions models conditional on the matching ratio, adjusting for unbalanced variables after PSM in subgroup analyses as follows: African American patients: The covariates of cohort month/year, age, ethnicity, insurance, medical history of autoimmune arthritis/rheumatoid arthritis, COPD, ILD, hypertension, HIV infection, smoking, diabetes, cancer, sickle cell disease, emergency room visits, and telemedicine visits were adjusted in the model. ≥65-year-old patients: The covariates of age (continuous), hypertension, and chronic kidney disease were adjusted in the model. <65-year-old patients: The covariates of cohort month/year and medical history of autoimmune arthritis/rheumatoid arthritis were adjusted in the model. The database may not completely collect vaccination data, which precluded assessing effectiveness in the unvaccinated patient subgroup. Error bars depict 95% CIs. PSM=propensity score matching.

## Discussion

In this real-world retrospective cohort study of US patients, the 30-day hospitalization incidence (95% CI) for high-risk COVID-19 patients prescribed nirmatrelvir/ritonavir was 1.21% (0.84−1.69) compared with 6.94% (6.03−7.94) for those who were not, corresponding to an 84% relative risk reduction in the nirmatrelvir/ritonavir group. The relative risk reduction for hospitalization within 15 days was 89%. These real-world data supporting nirmatrelvir/ritonavir effectiveness in decreasing hospitalization risk are extremely important given the continued global health threat of COVID-19, with Omicron-associated hospitalizations continuing to strain healthcare systems.^1, 21, 22^

In our subgroup analysis, nirmatrelvir/ritonavir prescription among African American patients was associated with a 65% relative risk reduction in 30-day hospitalization. The disparity between African American and White patients for hospitalization persisted among those prescribed nirmatrelvir/ritonavir with higher risk among African American patients, although 95% CIs were wide in the African American subgroup because of low number of events and sample size. Before PSM, African American patients comprised 5.9% of patients prescribed nirmatrelvir/ritonavir versus 13.5% of those who were not; respective percentages for White patients were 84.8% and 73.4%, indicating less frequent prescription of nirmatrelvir/ritonavir to African American patients. Our findings are consistent with established evidence of disproportionately higher COVID-19−associated hospitalization rates and healthcare disparities among African American patients.^12, 23, 24^ A clear and urgent need exists, therefore, to reduce such disparities among African American patients, including increasing access to COVID-19 antivirals.^12^ Adjusting high-risk eligibility criteria for African American patients so that more receive oral antivirals may address the hospitalization gap between African American and White patients observed in the nirmatrelvir/ritonavir group. Effectiveness among ≥65-year-olds was consistent with overall results and with those of younger patients, which is particularly important given clear evidence of increased hospitalization rates and worse COVID-19 outcomes among older patients.^14, 15, 25^

Our results support those from the EPIC-HR clinical trial, in which the relative risk of COVID-19−related hospitalization or all-cause death within 28 days among patients with similar high-risk characteristics or conditions who commenced treatment within 3 or 5 days of symptom onset was reduced by 87.8%−88.9% versus placebo.^4^ This is encouraging given the SARS-CoV-2 epidemiologic landscape differed between the 2 studies in that EPIC-HR was conducted during the Delta wave whereas the current study was conducted during Omicron predominance. In contrast to EPIC-HR, this real-world study did not use a composite effectiveness endpoint that included death because the exact day of death was not captured in the EHR database (only month of death was available^13^). Therefore, accurate characterization of nirmatrelvir/ritonavir effectiveness against mortality was not possible. This lack of precision led to the concern of an increased outcome misclassification and precluded an analysis of mortality taking a person-time approach. Mortality was therefore not included within the effectiveness endpoint, but was summarized as described to show potential censoring of hospitalizations. Our study also differed from EPIC-HR in that patients in this real-world analysis were permitted to have received COVID-19 vaccines (66%−68% of patients after PSM) or could have had previous SARS-CoV-2 infection (11%−12%), giving a broader picture of real-world use not obtainable from the clinical trial. The reduction in hospitalization risk among vaccinated patients underscores the concept that antivirals can complement vaccination in reducing the detrimental impact of COVID-19.^26^

Unlike EPIC-HR, which excluded patients with anticipated need for hospitalization within 48 hours after randomization,^4^ our analyses excluded patients hospitalized within 30 days before index date, which likely covers the period from symptom onset to ≤5 days of COVID-19 diagnosis. Patient demographic and clinical characteristics at baseline including this 30-day window were well balanced. Furthermore, we included patients hospitalized on their index date. For the nirmatrelvir/ritonavir group this meant the day of nirmatrelvir/ritonavir prescription. For the non-nirmatrelvir/ritonavir group (who were assigned their index date by PTDM), this was within 0−5 days from COVID-19 diagnosis. This approach is appropriate because COVID-19 symptomatology leading to hospitalization is an acute event and nirmatrelvir/ritonavir has a rapid onset of action;^27, 28^ same day COVID-19 diagnosis and nirmatrelvir/ritonavir prescription and hospitalization is therefore plausible. Excluding these patients would not be reflective of real-world use and could introduce a selection bias, as shown in a US real-world study in which excluding COVID-19 diagnoses coinciding with admissions led to lower hospitalization rates versus the overall population.^29^ Our approach is reinforced by monthly hospitalization rate data reported by the US CDC of 7.0%, 5.9%, 8.5%, 8.5%, 4.6%, and 3.7% (excluding patients with missing or unknown hospitalization status) for December 2021−May 2022,^30^ which are broadly similar to those within our study (6.1%, 6.3%, 10.1%, 7.8%, 4.8% and 3.2%, respectively).

Available real-world nirmatrelvir/ritonavir effectiveness studies conducted in the US (Massachusetts and New Hampshire) and Israel during the Omicron period identified 0.4%−0.7% hospitalization rates within 14−35 days among older (ie, ≥40-year-old or ≥50-year-old) patients who received nirmatrelvir/ritonavir compared with 1.0%−1.9% among those who did not.^29, 31^ An additional descriptive study found that <1% of US patients who received nirmatrelvir/ritonavir were hospitalized or visited the emergency department in the 5−15 days following treatment.^32^ By contrast, a Hong Kong study identified 4.4% and 6.2% hospitalization rates among nirmatrelvir/ritonavir and non-nirmatrelvir/ritonavir recipients, respectively, within 28 days of diagnosis.^33^ Our study found higher hospitalization rates than in the Massachusetts/New Hampshire and Israeli studies (nirmatrelvir/ritonavir group, 1.2%; non-nirmatrelvir/ritonavir group, 6.9%) and slightly lower rates than online-published results from a US database study (nirmatrelvir/ritonavir group, 1.9%; non-nirmatrelvir/ritonavir group, 9.7%^34^). Differences in hospitalization rates and effectiveness may be partially explained by differing percentages of patients who were vaccinated or had prior SARS-CoV-2 infection as well as differences in study design.^29, 31, 33, 35^ Additionally, 2 studies excluded patients whose initial diagnosis coincided with hospitalization admission, which may have biased hospitalization rates downward.^29, 35^ SARS-CoV-2 circulation and community immunity may also vary during different study periods. Despite variations in study methodology and effectiveness estimates, these real-world studies along with the current study collectively support nirmatrelvir/ritonavir effectiveness against Omicron-associated adverse outcomes.^29, 31-33, 35^

Study strengths include the large population-based design, which extracted data from a national longitudinal US EHR database and captured medical records linked by outpatients and inpatients within integrated delivery networks. We also employed various strategies to reduce effects of confounding variables or other biases that were not used, or were less comprehensive, in other nirmatrelvir/ritonavir real-world studies.^29, 31, 33, 35^ The PSM strategy used in our analysis to control for residual confounding was extensive and included covariates such as healthcare resource utilization, previous COVID-19 infection, and 50 most common medications, diagnoses, and procedures not often used in other real-world studies. Relatedly, patients in the nirmatrelvir/ritonavir and non-nirmatrelvir ritonavir groups were matched by cohort entry data to help reduce prescriber bias and variation in COVID-19−related hospitalization rates over time. We also used PTDM to control for immortal time bias, which arises in epidemiology studies using existing data, whereby there is a span of cohort follow-up when the studied outcome could not have occurred because of the exposure definition.^17^

Our study has several limitations. Optum data are encounter-based and do not cover all COVID-19 treatment and prescribing sites (eg, pharmacy, telehealth, public health departments). Thus, study patients in both groups can be sicker than the general population and a slower uptake of nirmatrelvir/ritonavir was observed. However, we expect that our cohort likely reflects the target population who are at higher risk for severe COVID-19 outcomes. Lack of medical chart review prevented effectively addressing exposure and outcome misclassifications. The database may not completely collect vaccination data, which precluded assessing effectiveness in the unvaccinated patient subgroup. We used hospitalization as a proxy of COVID-19−attributed hospitalization; however, because the data do not capture causes of hospitalization, inclusion of admission and encounters for conditions other than COVID-19 could result in outcome misclassification. The sensitivity analysis used a narrower risk window (ie, 15 days), as adopted in other real-world studies,^29, 32^ to help reduce potential effects of misclassification due to a longer risk window. However, this would not affect incidental hospitalization on the COVID-19 diagnosis date. There is a need to understand the frequency of non-COVID-19 hospitalizations carrying a COVID-19 diagnosis (specifically whether those are incident or past diagnoses). Nevertheless, hospitalizations occurring outside of integrated delivery networks would not be captured in the database, leading to potential underestimation of hospitalization in both groups. The EHR data source lacked the specific date of COVID-19 symptom onset, although this likely occurred ≤14 days from diagnosis for most cases.^36^ Nirmatrelvir/ritonavir effectiveness may also be underestimated because of incomplete treatment initiation, non-adherence, and undocumented nirmatrelvir/ritonavir receipt in the non-prescribed group. As previously mentioned, while death was included as a censoring event to define follow-up end date via an algorithm, mortality was not included as an endpoint because of concern of increased outcome misclassification. The study started when nirmatrelvir/ritonavir was granted EUA; its use has since expanded rapidly^34^ and earlier results may not reflect current use. Finally, study results may lack generalizability to patients not at high risk of severe disease, to other countries with different healthcare systems, and in non-US populations. Future research will aim to confirm these early real-world results, expand generalizability, and address limitations. We believe our findings, showing reduced COVID-19−associated hospitalization among those at high risk of severe disease, will assist prescribers and decision-makers to prioritize nirmatrelvir/ritonavir in vulnerable patient groups.

## Conclusions

Results from this study show that after proper adjustment for confounding, the real-world effectiveness of nirmatrelvir/ritonavir against hospitalization using a high-risk cohort during Omicron predominance supports the efficacy demonstrated in the EPIC-HR phase 3 trial.^4^ Nirmatrelvir/ritonavir was effective among vaccinated patients, and among White and African American patients, although prescribed less frequently to African American patients.

## Data Availability

Upon request, and subject to review, Pfizer will provide the summary data that support the findings of this study.

## Acknowledgments

The authors thank Sengwee Toh, ScD (Harvard Medical School and Harvard Pilgrim Health Care Institute) for invaluable discussions in the study design and development of the manuscript and Haitao Chu, MD, PhD (Pfizer Inc) for guidance regarding the study design and methods. Editorial/medical writing support was provided by Judith Kandel, PhD, Sheena Hunt, PhD, and Tricia Newell, PhD, at ICON (Blue Bell, PA, USA), and was funded by Pfizer Inc.

## References

1 World Health Organization. WHO Director-General’s opening remarks at the COVID-19 media briefing– 12 July 2022. https://www.who.int/director-general/speeches/detail/who-director-general-s-opening-remarks-at-the-covid-19-media-briefing--12-july-2022 (Accessed July 20 2022).

2 Center for Systems Science and Engineering at Johns Hopkins University. COVID-19 Dashboard. https://coronavirus.jhu.edu/map.html (Accessed July 15 2022).

3 Owen DR, Allerton CMN, Anderson AS, et al. An oral SARS-CoV-2 M(pro) inhibitor clinical candidate for the treatment of COVID-19. Science 2021;374(6575):1586–93. doi:10.1126/science.abl4784

4 Hammond J, Leister-Tebbe H, Gardner A, et al. Oral Nirmatrelvir for High-Risk, Nonhospitalized Adults with Covid-19. N Engl J Med 2022;386(15):1397–408. doi:10.1056/NEJMoa2118542

5 Pfizer Inc. Fact sheet for healthcare providers: Emergency use authorization for PAXLOVID™ 2022.

6 Sherman RE, Anderson SA, Dal Pan GJ, et al. Real-World Evidence - What Is It and What Can It Tell Us? N Engl J Med 2016;375(23):2293–97. doi:10.1056/NEJMsb1609216

7 Htoo PT, Measer G, Orr R, et al. Evaluating Confounding Control in Estimations of Influenza Antiviral Effectiveness in Electronic Health Plan Data. Am J Epidemiol 2022;191(5):908–20. doi:10.1093/aje/kwac020

8 Norgaard M, Ehrenstein V, Vandenbroucke JP. Confounding in observational studies based on large health care databases: problems and potential solutions - a primer for the clinician. Clin Epidemiol 2017;9:185–93. doi:10.2147/CLEP.S129879

9 US National Institutes of Health. Ritonavir-Boosted Nirmatrelvir (Paxlovid). 2022. https://www.covid19treatmentguidelines.nih.gov/therapies/antiviral-therapy/ritonavir-boosted-nirmatrelvir--paxlovid-/ (Accessed July 20 2022).

10 Dal-Re R, Becker SL, Bottieau E, et al. Availability of oral antivirals against SARS-CoV-2 infection and the requirement for an ethical prescribing approach. Lancet Infect Dis 2022;22(8):e231–e38. doi:10.1016/S1473-3099(22)00119-0

11 Hodcroft EB. CoVariants: SARS-CoV-2 Mutations and Variants of Interest. 2022. https://covariants.org/ (Accessed January 7 2022).

12 Mude W, Oguoma VM, Nyanhanda T, et al. Racial disparities in COVID-19 pandemic cases, hospitalisations, and deaths: A systematic review and meta-analysis. J Glob Health 2021;11:05015. doi:10.7189/jogh.11.05015

13 Page JH, Londhe AA, Brooks C, et al. Trends in characteristics and outcomes among US adults hospitalised with COVID-19 throughout 2020: an observational cohort study. BMJ Open 2022;12(2):e055137. doi:10.1136/bmjopen-2021-055137

14 Li X, Zhong X, Wang Y, et al. Clinical determinants of the severity of COVID-19: A systematic review and meta-analysis. PLoS One 2021;16(5):e0250602. doi:10.1371/journal.pone.0250602

15 Docherty AB, Harrison EM, Green CA, et al. Features of 20 133 UK patients in hospital with covid-19 using the ISARIC WHO Clinical Characterisation Protocol: prospective observational cohort study. BMJ 2020;369:m1985. doi:10.1136/bmj.m1985

16 Yang J, Ma Z, Lei Y. A meta-analysis of the association between obesity and COVID-19. Epidemiol Infect 2020;149:e11. doi:10.1017/S0950268820003027

17 Levesque LE, Hanley JA, Kezouh A, et al. Problem of immortal time bias in cohort studies: example using statins for preventing progression of diabetes. BMJ 2010;340:b5087. doi:10.1136/bmj.b5087

18 Zhou Z, Rahme E, Abrahamowicz M, et al. Survival bias associated with time-to-treatment initiation in drug effectiveness evaluation: a comparison of methods. Am J Epidemiol 2005;162(10):1016–23. doi:10.1093/aje/kwi307

19 Austin PC. An Introduction to Propensity Score Methods for Reducing the Effects of Confounding in Observational Studies. Multivariate Behav Res 2011;46(3):399–424. doi:10.1080/00273171.2011.568786

20 Zhang Z, Kim HJ, Lonjon G, et al. Balance diagnostics after propensity score matching. Ann Transl Med 2019;7(1):16. doi:10.21037/atm.2018.12.10

21 Iuliano AD, Brunkard JM, Boehmer TK, et al. Trends in Disease Severity and Health Care Utilization During the Early Omicron Variant Period Compared with Previous SARS-CoV-2 High Transmission Periods - United States, December 2020-January 2022. MMWR Morb Mortal Wkly Rep 2022;71(4):146–52. doi:10.15585/mmwr.mm7104e4

22 Myers LC, Liu VX. The COVID-19 Pandemic Strikes Again and Again and Again. JAMA Netw Open 2022;5(3):e221760. doi:10.1001/jamanetworkopen.2022.1760

23 Andraska EA, Alabi O, Dorsey C, et al. Health care disparities during the COVID-19 pandemic. Semin Vasc Surg 2021;34(3):82–88. doi:10.1053/j.semvascsurg.2021.08.002

24 Tai DBG, Shah A, Doubeni CA, et al. The Disproportionate Impact of COVID-19 on Racial and Ethnic Minorities in the United States. Clin Infect Dis 2021;72(4):703–06. doi:10.1093/cid/ciaa815

25 Ko JY, Danielson ML, Town M, et al. Risk Factors for Coronavirus Disease 2019 (COVID-19)-Associated Hospitalization: COVID-19-Associated Hospitalization Surveillance Network and Behavioral Risk Factor Surveillance System. Clin Infect Dis 2021;72(11):e695–e703. doi:10.1093/cid/ciaa1419

26 Matrajt L, Brown ER, Cohen MS, et al. Could widespread use of antiviral treatment curb the COVID-19 pandemic? A modeling study. medRxiv 2022 doi:10.1101/2021.11.10.21266139

27 Singh RSP, Toussi SS, Hackman F, et al. Innovative Randomized Phase I Study and Dosing Regimen Selection to Accelerate and Inform Pivotal COVID-19 Trial of Nirmatrelvir. Clin Pharmacol Ther 2022;112(1):101–11. doi:10.1002/cpt.2603

28 Toussi SS, Neutel JM, Navarro J, et al. Pharmacokinetics of Oral Nirmatrelvir/Ritonavir, a Protease Inhibitor for Treatment of COVID-19, in Subjects With Renal Impairment. Clin Pharmacol Ther 2022 doi:10.1002/cpt.2688

29 Dryden-Peterson S, Kim A, Kim AY, et al. Nirmatrelvir plus ritonavir for early COVID-19 and hospitalization in a large US health system. medRxiv 2022 doi:10.1101/2022.06.14.22276393

30 US Centers for Disease Control and Prevention. COVID-19 Case Surveillance Public Use Data with Geography. 2022. https://data.cdc.gov/Case-Surveillance/COVID-19-Case-Surveillance-Public-Use-Data-with-Ge/n8mc-b4w4 (Accessed July 22 2022).

31 Arbel R, Wolff Sagy Y, Hoshen M, et al. Nirmatrelvir Use and Severe Covid-19 Outcomes during the Omicron Surge. N Engl J Med 2022;387(9):790–98. doi:10.1056/NEJMoa2204919

32 Malden DE, Hong V, Lewin BJ, et al. Hospitalization and Emergency Department Encounters for COVID-19 After Paxlovid Treatment - California, December 2021-May 2022. MMWR Morb Mortal Wkly Rep 2022;71(25):830–33. doi:10.15585/mmwr.mm7125e2

33 Wong CKH, Au ICH, Lau KTK, et al. Real-world effectiveness of molnupiravir and nirmatrelvir/ritonavir against mortality, hospitalization, and in-hospital outcomes among community-dwelling, ambulatory COVID-19 patients during the BA.2.2 wave in Hong Kong: an observational study. medRxiv 2022 doi:10.1101/2022.05.26.22275631

34 Allen S, Johnson K, Haddock J, et al. Game Changer: Paxlovid Reduces Hospitalizations and Saves Lives. 2022. https://epicresearch.org/articles/game-changer-paxlovid-reduces-hospitalizations-and-saves-lives (Accessed August 3 2022).

35 Najjar-Debbiny R, Gronich N, Weber G, et al. Effectiveness of Paxlovid in Reducing Severe COVID-19 and Mortality in High Risk Patients. Clin Infect Dis 2022 doi:10.1093/cid/ciac443

36 Griffin DO, Brennan-Rieder D, Ngo B, et al. The Importance of Understanding the Stages of COVID-19 in Treatment and Trials. AIDS Rev 2021;23(1):40–47. doi:10.24875/AIDSRev.200001261

