## Supplementary material for "Real-World Effectiveness of Nirmatrelvir/Ritonavir in Preventing Hospitalization Among Patients With COVID-19 at High Risk for Severe Disease in the United States: A Nationwide Population-Based Cohort Study"

### Supplementary Appendix

### Optum® De-Identified COVID-19 Electronic Health Record Dataset

The Optum de-identified COVID-19 Electronic Health Record (EHR) dataset is a subset of the Optum EHR data, which is reflective of the demographic distribution by age and sex of the US population.[1,2] The database is certified as de-identified and is therefore exempt from institutional review board approval.[2] De-identified clinical data from numerous health plans, providers, and payers (ie, public, commercial, other types, uninsured) are included, which is integrated across care settings and are longitudinally linked by patient.[1,2] Inclusion of data from 2007 in the Optum COVID-19 EHR database allows analysis of patient medical history.[1] A limitation of the Optum COVID-19 EHR database is that it may not capture records outside of the integrated delivery network and death events are recorded at the month level, to reduce the risk of patient identifiability.[2]

### Characteristics or Underlying Medical Conditions Associated with Increased Risk of Developing Severe COVID-19

Patients had to have ≥1 of the following characteristics or underlying medical conditions associated with increased risk of developing severe COVID-19 at baseline unless otherwise specified: age ≥60 years at index date; body mass index >25 kg/m$^2$; current smoker; immunosuppressive disease or prolonged use of immune-weakening medications; chronic lung disease, hypertension, cardiovascular disease, diabetes mellitus, chronic kidney disease, sickle cell disease, neurodevelopmental disorders or other medically complex conditions; or cancer diagnosis (other than localized skin cancer) ≤1 year before the index date. Patients were excluded if they had any of the following high-risk criteria at any time before the index date (unless specified otherwise): known medical history of active liver disease (other than nonalcoholic hepatic steatosis), including chronic or active hepatitis B or C infection, primary biliary cirrhosis, or acute liver failure; dialysis receipt or end stage of renal disease; or pregnant or lactating within 9 months before index date.

[1] Curtis JR et al. J Rheumatol. 2022;49:320-329.
[2] Page JH et al. BMJ Open. 2022;12:e055137.

### COVID-19 Treatments or Contraindicated Medications

Patients in both groups were excluded if they had taken any of the following medications within 28 days of index date:

| COVID-19 treatments in use or recommended at the time of the study[3] | Contraindicated medications | |
|---|---|---|
| **Anti-SARS-CoV-2 antibody products** | **Alpha1-adrenoreceptor antagonist** | **HMG-CoA reductase inhibitors** |
| Bamlanivimab/etesevimab | Alfuzosin | Lovastatin |
| Casirivimab/imdevimab | **Analgesics** | Simvastatin |
| Sotrovimab | Pethidine | **PDE5 inhibitor** |
| Tixagevimab/cilgavimab | Propoxyphene | Sildenafil |
| Bebtelovimab | **Antianginal** | **Sedative/hypnotics** |
| **Convalescent plasma** | Ranolazine | Triazolam |
| **Antiviral therapy** | **Antiarrhythmic** | Oral midazolam |
| Remdesivir | Amiodarone | **Anticancer drugs** |
| Molnupiravir | Dronedarone | Apalutamide |
| Chloroquine/hydroxychloroquine | Flecainide | **Anticonvulsant** |
| Azithromycin | Propafenone | Carbamazepine |
| Lopinavir/ritonavir | Quinidine | Phenobarbital |
| Ivermectin | **Anti-gout** | Phenytoin |
| Nitazoxanide | Colchicine | **Herbal products** |
| Ribavirin | **Antipsychotics** | St. John's Wort |
| Interferon-beta | Clozapine | |
| **Immunomodulators** | Pimozide | |
| **Systemic corticosteroids** | Lurasidone | |
| Dexamethasone | **Interleukin inhibitors** | |
| Methylprednisolone | Tocilizumab | |
| **Fluvoxamine** | Sarilumab | |
| **Antimycobacterials** | **Ergot derivatives** | |
| Rifampin | Ergotamine | |
| **Antithrombotic therapy** | Methylergonovine | |
| Heparin | Dihydroergotamine | |

[3] NIH Coronavirus Disease 2019 (COVID-19) Treatment Guidelines. Last update: April 1, 2022. Accessed on: April 24, 2022. https://files.covid19treatmentguidelines.nih.gov/guidelines/archive/covid19treatmentguidelines-04-01-2022.pdf.

### Prescription Time Distribution Method

Index dates were assigned to individuals in the non-nirmatrelvir/ritonavir group using the Prescription Time Distribution Method (PTDM)[4] as follows. Among all patients in the nirmatrelvir/ritonavir group with a date of confirmed COVID-19 diagnosis, the time from the date of confirmed COVID-19 to the date of nirmatrelvir/ritonavir prescription was calculated. The percentages of those prescribed nirmatrelvir/ritonavir on Day 0 (date of COVID-19 diagnosis), and Days 1, 2, 3, and 4 were calculated. Patients in the non-nirmatrelvir/ritonavir group were randomly assigned index dates relative to their COVID-19 diagnosis dates based on the resulting distribution of the index date in the nirmatrelvir/ritonavir group. For example, if 20% of nirmatrelvir/ritonavir patients were prescribed nirmatrelvir/ritonavir on Day 1 relative to their confirmed COVID-19 diagnosis, then 20% of non-nirmatrelvir/ritonavir patients were randomly selected and assigned their index date to be the date of COVID-19 diagnosis plus 1 day.

### Algorithm for Assigning Date of Death

In the Optum database, deaths are recorded by month and year and not by exact date. The following algorithm was applied to assign a death date to each patient who died during follow-up:

1) Assign 15th day of the month as the death date;
2) If death occurred before the index date but on the same month, assign a death date of the last day of the month of death;
3) If death occurred after the end of follow-up but on the same month, assign the death date to be the same as the follow-up end date and consider this patient to have died in the study.

### Loss to Follow-up

For any patient in the database, the last active date was derived from the last date of any healthcare activity (eg, outpatient, inpatient, emergency room, prescription, laboratory value). If a patient had the last active date before the end of the 30-day follow-up window (primary analysis), without hospitalization, this patient was considered lost to follow-up in the analysis. Overall, 1339 (48%) and 4686 (43%) patients in the nirmatrelvir/ritonavir and non-nirmatrelvir/ritonavir groups, respectively, had their last active date before the end of the 30-day window and were considered lost to follow-up.

[4] Zhou Z et al. Am J Epidemiol 2005;162:1016-1023.

Corresponding values before the end of the 15-day window were 938 (33%) and 3345 (31%) patients in the nirmatrelvir/ritonavir and non-nirmatrelvir/ritonavir groups, respectively. Since Optum COVID-19 EHR data are encounter-based and the study period is short (December 22, 2022 to June 8, 2022) loss to follow-up during the 30-day risk window for a patient shows that these patients had no additional healthcare utilization during those 30 days. It does not necessarily mean that this patient left the healthcare system; thus, the proportion of those lost to follow-up should be interpreted with caution.

### Table S1. Healthcare utilization within 12 months before index date before and after PSM

| | Before PSM | | | After PSM | | |
|---|---|---|---|---|---|---|
| | Nirmatrelvir/ Ritonavir N=2811 | Non-Nirmatrelvir/ Ritonavir N=194,542 | Standardized Difference | Nirmatrelvir/ Ritonavir N=2808 | Non-Nirmatrelvir/ Ritonavir N=10,849 | Standardized Difference |
| Number of hospitalizations | | | | | | |
| 0 | 2624 (93.3) | 179,547 (92.3) | 0.041 | 2621 (93.3) | 10,123 (93.3) | 0.001 |
| 1 | 154 (5.5) | 11,748 (6.0) | 0.024 | 154 (5.5) | 587 (5.4) | 0.003 |
| ≥2 | 33 (1.2) | 3247 (1.7) | 0.042 | 33 (1.2) | 139 (1.3) | 0.009 |
| Mean ± standard deviation | 0.08 ± 0.34 | 0.10 ± 0.42 | 0.053 | 0.08 ± 0.34 | 0.09 ± 0.38 | 0.014 |
| Length of stay, days | | | | | | |
| Median, IQR | 4.00, 7.00 | 4.00, 6.00 | - | 4.00, 7.00 | 4.00, 6.00 | - |
| Emergency room visits | | | | | | |
| 0 | 2357 (83.8) | 136,186 (70.0) | 0.333 | 2354 (83.8) | 9094 (83.8) | 0.004 |
| 1 | 286 (10.2) | 31,146 (16.0) | 0.174 | 286 (10.2) | 1130 (10.4) | 0.005 |
| ≥2 | 168 (6.0) | 27,210 (14.0) | 0.270 | 168 (6.0) | 625 (5.8) | 0.013 |
| Mean ± standard deviation | 0.27 ± 0.79 | 0.65 ± 1.97 | 0.252 | 0.27 ± 0.79 | 0.29 ± 0.94 | 0.023 |
| Physician office visits | | | | | | |
| 0 | 265 (9.4) | 29,125 (15.0) | 0.170 | 265 (9.4) | 1058 (9.8) | 0.002 |
| 1 | 335 (11.9) | 23,626 (12.1) | 0.007 | 335 (11.9) | 1269 (11.7) | 0.012 |
| ≥2 | 2211 (78.7) | 141,791 (72.9) | 0.135 | 2208 (78.6) | 8522 (78.6) | 0.008 |

| | | | | | | |
|---|---|---|---|---|---|---|
| Mean ± standard deviation | 6.78 ± 8.07 | 5.88 ± 7.74 | 0.113 | 6.77 ± 8.07 | 6.74 ± 7.92 | 0.008 |
| Telemedicine visits | | | | | | |
| 0 | 1287 (45.8) | 95,571 (49.1) | 0.067 | 1287 (45.8) | 5023 (46.3) | 0.006 |
| 1 | 417 (14.8) | 28,119 (14.5) | 0.011 | 415 (14.8) | 1561 (14.4) | 0.012 |
| ≥2 | 1107 (39.4) | 70,852 (36.4) | 0.061 | 1106 (39.4) | 4265 (39.3) | 0.002 |
| Mean ± standard deviation | 2.79 ± 5.75 | 2.39 ± 4.88 | 0.074 | 2.79 ± 5.75 | 2.71 ± 5.25 | 0.003 |
| Urgent care visits | | | | | | |
| 0 | 2653 (94.4) | 171,289 (88.0) | 0.225 | 2650 (94.4) | 10,225 (94.2) | 0.001 |
| 1 | 98 (3.5) | 13,321 (6.8) | 0.152 | 98 (3.5) | 385 (3.5) | 0.001 |
| ≥2 | 60 (2.1) | 9932 (5.1) | 0.160 | 60 (2.1) | 239 (2.2) | 0.000 |
| Mean ± standard deviation | 0.10 ± 0.57 | 0.23 ± 0.88 | 0.175 | 0.10 ± 0.57 | 0.10 ± 0.52 | 0.016 |

IQR, interquartile range; PSM=propensity score matching.

Values shown are n (%) unless otherwise specified.

**Table S2. Baseline clinical characteristics before and after PSM reported in <5% in both groups**

| | Before PSM | | | After PSM | | |
|---|---|---|---|---|---|---|
| **Comorbidity, n (%)** | **Nirmatrelvir/ Ritonavir N=2811** | **Non-Nirmatrelvir/ Ritonavir N=194,542** | **Standardized Difference** | **Nirmatrelvir/ Ritonavir N=2808** | **Non-Nirmatrelvir/ Ritonavir N=10,849** | **Standardized Difference** |
| Multiple sclerosis | 15 (0.5) | 786 (0.4) | 0.019 | 15 (0.5) | 54 (0.5) | 0.005 |
| Sjögren's syndrome | 19 (0.7) | 615 (0.3) | 0.051 | 19 (0.7) | 52 (0.5) | 0.025 |
| Guillain-Barré | 1 (0.0) | 51 (0.0) | 0.005 | 1 (0.0) | 6 (0.1) | 0.008 |
| Idiopathic thrombocytopenia purpura | 0 (0.0) | 175 (0.1) | - | 0 (0.0) | 10 (0.1) | - |
| Autoimmune thyroiditis | 43 (1.5) | 1763 (0.9) | 0.057 | 43 (1.5) | 127 (1.2) | 0.031 |
| Giant cell arteritis | 0 (0.0) | 56 (0.0) | - | 0 (0.0) | 8 (0.1) | - |
| Psoriasis | 62 (2.2) | 1920 (1.0) | 0.097 | 62 (2.2) | 153 (1.4) | 0.057 |
| Systemic or cutaneous lupus erythematosus | 29 (1.0) | 892 (0.5) | 0.067 | 29 (1.0) | 60 (0.6) | 0.054 |
| Autoimmune arthritis/rheumatoid arthritis | 73 (2.6) | 2506 (1.3) | 0.095 | 73 (2.6) | 211 (1.9) | 0.043 |
| Interstitial lung disease | 43 (1.5) | 1400 (0.7) | 0.077 | 43 (1.5) | 127 (1.2) | 0.032 |
| Cardiomyopathies | 50 (1.8) | 2633 (1.4) | 0.034 | 50 (1.8) | 209 (1.9) | 0.012 |

| | | | | | | |
|---|---|---|---|---|---|---|
| Cerebrovascular disease or stroke (including transient ischemic attack) | 130 (4.6) | 6415 (3.3) | 0.068 | 130 (4.6) | 529 (4.9) | 0.015 |
| HIV infection | 20 (0.7) | 701 (0.4) | 0.048 | 20 (0.7) | 41 (0.4) | 0.047 |
| Sickle cell disease | 4 (0.1) | 341 (0.2) | 0.008 | 4 (0.1) | 3 (0.0) | 0.038 |

PSM=propensity score matching.

Values shown are n (%).

**Table S3. Hospitalization among subgroups of patients with COVID-19 by nirmatrelvir/ritonavir prescription status after PSM**

| | Hospitalization within 30 days | | Hospitalization within 15 days | |
|---|---|---|---|---|
| | Nirmatrelvir/Ritonavir | Non-Nirmatrelvir/Ritonavir | Nirmatrelvir/Ritonavir | Non-Nirmatrelvir/Ritonavir |
| Race | | | | |
| White | | | | |
| Patients at risk | 2381 | 9132 | 2381 | 9132 |
| Events | 24 | 634 | 14 | 595 |
| Incidence proportion, % (95% CI) | 1.01 (0.65, 1.50) | 6.94 (6.43, 7.48) | 0.59 (0.32, 0.98) | 6.52 (6.02, 7.04) |
| Person-days | 47,277 | 172,909 | 26,844 | 97,181 |
| Incidence rate, per 100 person-days (95% CI) | 0.05 (0.03, 0.08) | 0.37 (0.34, 0.40) | 0.05, (0.03, 0.09) | 0.61 (0.56, 0.66) |
| Hazard ratio (95% CI) | 0.14 (0.09, 0.21) | - | 0.09 (0.05, 0.15) | - |
| Relative risk reduction, % (95% CI) | 86 (79, 81) | - | 91 (85, 95) | - |
| African American[a] | | | | |
| Patients at risk | 167 | 662 | 167 | 662 |
| Events | 8 | 58 | 7 | 56 |
| Incidence proportion, % (95% CI) | 4.79 (2.09, 9.22) | 8.76 (6.72, 11.18) | 4.19 (1.70, 8.45) | 8.46 (6.45, 10.84) |
| Person-days | 3504 | 11,813 | 1987 | 6653 |
| Incidence rate, per 100 person-days (95% CI) | 0.23 (0.10, 0.45) | 0.49 (0.37, 0.63) | 0.35 (0.14, 0.73) | 0.84 (0.64, 1.09) |

| | | | | |
|---|---|---|---|---|
| Hazard ratio (95% CI) | 0.35 (0.15, 0.83) | - | 0.33 (0.13, 0.83) | - |
| Relative risk reduction, % (95% CI) | 65 (17, 85) | - | 67 (17, 87) | - |
| Age | | | | |
| ≥65 years[b] | | | | |
| Patients at risk | 1250 | 4723 | 1250 | 4723 |
| Events | 26 | 583 | 16 | 555 |
| Incidence proportion, % (95% CI) | 2.08 (1.36, 3.03) | 12.34 (11.42, 13.32) | 1.28 (0.73, 2.07) | 11.75 (10.85, 12.70) |
| Person-days | 25,399 | 84,908 | 14,315 | 48,069 |
| Incidence rate, per 100 person-days (95% CI) | 0.10 (0.07, 0.15) | 0.69 (0.63, 0.74) | 0.11 (0.06, 0.18) | 1.16 (1.06, 1.25) |
| Hazard ratio (95% CI) | 0.17 (0.12, 0.26) | - | 0.11 (0.07, 0.19) | - |
| Relative risk reduction, % (95% CI) | 83 (74, 88) | - | 89 (91, 93) | - |
| <65 years[c] | | | | |
| Patients at risk | 1558 | 6126 | 1558 | 6126 |
| Events | 8 | 169 | 6 | 153 |
| Incidence proportion, % (95% CI) | 0.51 (0.22, 1.01) | 2.76 (2.36, 3.20) | 0.39 (0.14, 0.84) | 2.50 (2.12, 2.92) |
| Person-days | 30,562 | 120,072 | 17,406 | 67,027 |
| Incidence rate, per 100 person-days (95% CI) | 0.03 (0.01, 0.05) | 0.14 (0.12, 0.16) | 0.03 (0.01, 0.08) | 0.23 (0.19 0.27) |
| Hazard ratio (95% CI) | 0.19 (0.09, 0.38) | - | 0.16 (0.07, 0.35) | - |
| Relative risk reduction, % (95% CI) | 81 (62, 91) | - | 84 (65, 93) | - |

| | | | | |
|---|---|---|---|---|
| Vaccination status | | | | |
| Vaccinated | | | | |
| Patients at risk | 1897 | 7207 | 1897 | 7207 |
| Events | 20 | 412 | 11 | 380 |
| Incidence proportion, % (95% CI) | 1.05 (0.65, 1.62) | 5.72 (5.19, 6.28) | 0.58 (0.29, 1.04) | 5.27 (4.77, 5.81) |
| Person-days | 39,052 | 146,112 | 22,078 | 81,280 |
| Incidence rate, per 100 person-days (95% CI) | 0.05 (0.03, 0.08) | 0.28 (0.26, 0.31) | 0.05 (0.02, 0.09) | 0.47 (0.42, 0.52) |
| Hazard ratio (95% CI) | 0.18 (0.12, 0.28) | - | 0.11 (0.06, 0.20) | - |
| Relative risk reduction, % (95% CI) | 82 (72, 88) | - | 89 (80, 94) | - |

COPD=chronic obstruction pulmonary disease; ILD=interstitial lung disease; PSM=propensity score matching.

Hazard ratios and 95% CIs were computed using Cox proportional hazards regressions models. Relative risk reductions were derived by subtracting the hazard ratios from 1.

[a]The covariates of cohort month/year, age, ethnicity, insurance, medical history of autoimmune arthritis/rheumatoid arthritis, COPD, ILD, hypertension, HIV infection, smoking, diabetes, cancer, sickle cell disease, emergency room visits, and telemedicine visits were adjusted in the model among African American patients.

[b]The covariates of age (continuous), hypertension, and chronic kidney disease were adjusted in the model among ≥65-year-old patients.

[c]The covariates of cohort month/year and medical history of autoimmune arthritis/rheumatoid arthritis were adjusted in the model among <65-year-old patients.

### Figure S1. Hospitalization by month: all patients

Blue bars represent hospitalization rates for all Optum patients with COVID-19 meeting study eligibility criteria and having index dates between December 22, 2021 and May 8, 2022. Black bars represent published CDC COVID-19 hospitalization rates during the same time period.[5]

CDC=US Centers for Disease Control and Prevention.

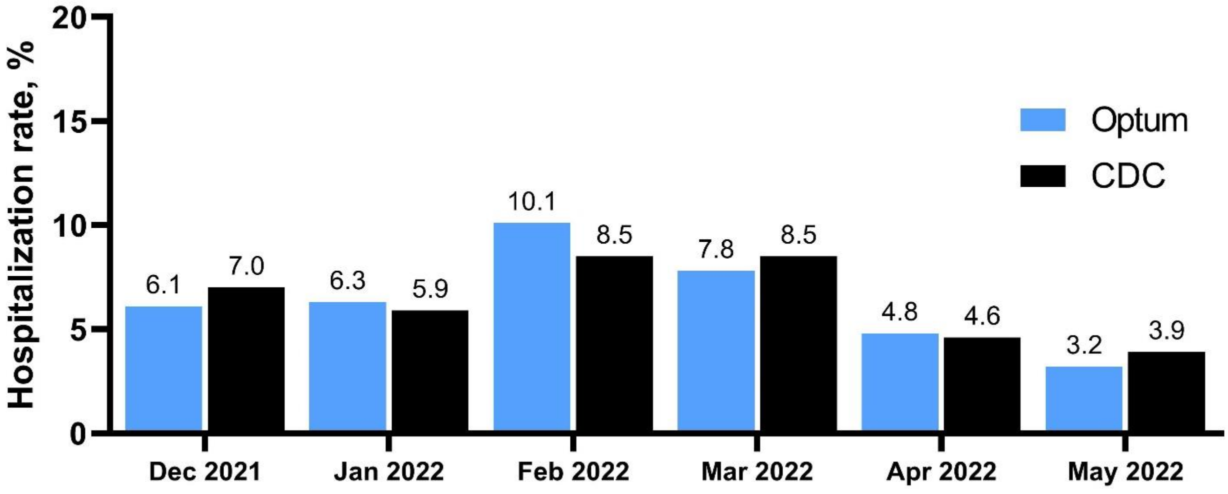


[5] US Centers for Disease Control and Prevention. COVID-19 Case Surveillance Public Use Data with Geography. Available at: https://data.cdc.gov/Case-Surveillance/COVID-19-Case-Surveillance-Public-Use-Data-with-Ge/n8mc-b4w4. Accessed July 22, 2022.
